# Characterisation of type 2 diabetes subgroups and their association with ethnicity and clinical outcomes: a UK real-world data study using the East London Database

**DOI:** 10.1101/2021.08.26.21262657

**Authors:** Rohini Mathur, Sally A Hull, Sam Hodgson, Sarah Finer

**Author notes:** Corresponding Author: Dr Rohini Mathur.

## Abstract

**Background:** Subgroups of type 2 diabetes (T2DM) have been well characterised in experimental studies. However, it is unclear whether T2DM subgroups can be identified in UK based real-world populations and if they impact clinical outcomes.

**Aim:** To derive T2DM subgroups using primary care data from a multi-ethnic population, evaluate associations with glycaemic control, treatment initiation and vascular outcomes, and understand how these vary by ethnicity.

**Design and setting:** An observational cohort study in the East London Primary Care Database from 2008-2018.

**Method:** Latent class analysis using age, sex, glycated haemoglobin, and body mass index at diagnosis was used to derive T2DM subgroups in White, South Asian, and Black groups. Time to treatment initiation and vascular outcomes was estimated using multivariable Cox-proportional hazards regression.

**Results:** 31,931 adults with T2DM were included: 47% south Asian, 25% White, 20% Black. We replicated two previously described subgroups, ‘Mild Age-Related Diabetes’ (MARD), ‘Mild Obesity-related Diabetes (MOD), and characterised a third ‘Severe Hyperglycaemic Diabetes’ (SHD). Compared to MARD, SHD had the poorest long term glycaemic control, fastest initiation of antidiabetic treatment (HR 2.02, 1.76-2.32), and highest risk of microvascular complications (HR 1.38, 1.28-1.49). MOD had the highest risk of macrovascular complications (HR 1.50, 1.23-1.83). Subgroup differences in treatment initiation were most pronounced for the White group, and vascular complications for the Black group.

**Conclusions:** Clinically useful T2DM subgroups, identified at diagnosis, can be generated in routine real-world multi-ethnic populations, and may offer a pragmatic means to develop stratified primary care pathways and improve healthcare resource allocation.

**How this fits in:** Previous studies of predominantly White European populations have identified four T2DM subgroups. In the UK the clinical measures necessary to replicate these subgroups are only available in secondary care data, limiting their usefulness for diabetes management in primary care settings. In this study, we demonstrate how clinically meaningful T2DM subgroups can be pragmatically generated using real-world primary care data. Furthermore, we highlight important differences between T2DM subgroups with respect to vascular outcomes, treatment initiation, and HbA1c control. Diabetes subgroups are a useful heuristic for helping clinician decision-making which, in turn, this can lead to a more personalised design of diabetes care focussed on more intensive management of subgroups most at risk of complications, such as those with severe hyperglycaemia at time of diagnosis.

## Introduction

Type 2 diabetes (T2DM) is a heterogenous, multifactorial condition with major impact on the health of global populations and economic cost, largely mediated through its vascular complications.^1,2^ Recent studies have identified distinct and replicable subgroups of T2DM in both experimental^3–5^ and non-experimental (real-world) cohorts using data-driven clustering methods.^6–8^ These studies have defined T2DM subgroups using clinical variables typically assessed in secondary care settings, including measures of insulin secretion and resistance (determined using C-peptide and glucose assays), to delineate subgroups and their likely aetiology.^3–5,8^ However, in the UK, the majority of type 2 diabetes management takes place in primary care settings, where these investigations are rarely performed. It is not known whether using data routinely available in the primary care setting can also be used to identify T2DM subgroups and their association with clinical outcomes. The impact of ethnicity on the characterisation T2DM subgroups is also poorly understood but is an important area of study given the varied aetiological processes and disease prevalence and outcomes seen in different ethnic groups with the condition.^9–11^

We therefore set out to characterise Type 2 diabetes subgroups in a large UK-based multi-ethnic population using routinely collected clinical measures captured in the primary care record. We then sought to investigate differences in control of HbA1c, time to initiation of antidiabetic treatment, and risk of vascular outcomes by diabetes subgroup and ethnicity. In doing so, we evaluate the potential utility of data-driven T2D subgroup identification to stratify care delivery in NHS primary care.

## Methods

### Study Population

An observational cohort study was conducted using the East London Primary Care Database, which includes anonymised longitudinal health record data from 1 million individuals registered at 125 general practices across the three multi-ethnic inner-London boroughs of Tower Hamlets, Newham, and Hackney.^12^ The study population included all adults aged 18 and over newly diagnosed with T2DM between January 2007 and January 2018 with at least 12 months of continuous registration prior to first diagnosis of type 2 diabetes. Type 2 diabetes diagnosis was identified using the C10F% Read codes as defined in the UK Quality and Outcomes Framework.^13^

### Covariates

Self-reported ethnicity was identified using Read codes and collapsed into the five high-level categories of the 2011 Census (White, South Asian, Black African/Caribbean, Mixed, and Other) (see supplementary material). Age at T2DM diagnosis was calculated from the date of diagnosis and age at data extraction (January 2018). Deprivation was measured using the Townsend score and divided into quintiles. Baseline was defined as the date of type 2 diagnosis and baseline measures of glycated haemoglobin (HbA1c), body mass index (BMI), systolic and diastolic blood pressure (SBP, DBP), total cholesterol, and estimated glomerular filtration rate (eGFR) were derived from the last value in the year prior to diagnosis. Comorbid conditions were considered prevalent at baseline if present on the clinical record at any date prior to type 2 diabetes diagnosis. These included hypertension, coronary heart disease (CHD), stroke, heart failure, chronic kidney disease (CKD), retinopathy, and neuropathy. Antidiabetic medications were categorized as all oral non-insulin antidiabetic drugs and insulin. Macrovascular disease was defined as a composite of coronary heart disease, heart failure, myocardial infarction, and stroke. Microvascular disease was defined as a composite of chronic kidney disease, neuropathy, and retinopathy. Incident macrovascular and microvascular events were defined as diagnoses recorded at any point after T2DM diagnosis.

### Statistical Analysis

Latent class analysis was used to identify subgroups of Type 2 diabetes for the whole cohort and separately for individuals of white, South Asian, and Black ethnicity. Subgroups were derived using data from four observed indicator variables: age at diagnosis, sex, HbA1c at diagnosis, and BMI at diagnosis. Models with between 2-5 classes were compared and the optimal number of classes was chosen by evaluating the Bayesian Information Criteria (with lower values indicating better fit), clinical interpretability, minimum posterior probability of group membership over 70%, and sufficient group membership, defined as >1% of the study population in each class.

Amongst those free from prevalent vascular disease at baseline, age-sex adjusted Cox-proportional hazards regression was used to compare the risk of incident macro- and microvascular disease between T2DM subgroups by ethnic group. Amongst individuals free from any antidiabetic treatment at diagnosis, differences in time to initiation of antidiabetic treatment between subgroups for the whole population, and by ethnic group were modelled using age-sex adjusted multivariable cox-proportional hazards regression. Individuals who initiated treatment in the 30 days prior to diagnosis were included in the analysis by moving their treatment initiation date to one day after T2DM diagnosis as they were considered to be ‘baseline initiators’, for whom the initial prescription formed part of the diabetes diagnosis process. HbA1c in the five years following initial diagnosis was calculated for each individual by taking the mean of all HbA1c values recorded in each 12-month period. Mean HbA1c in each ethnic group and latent class was then plotted over time.

## Results

A total of 31,931 adults with type 2 diabetes were included in the study, of whom 47% were of south Asian ethnicity (n=14,884), 25% were of white ethnicity (n=8,154), 20% were of black ethnicity (n=6,423) and 6% were of mixed or other ethnicities (n=1,957). Ethnicity was unknown for 1.6% of the study population (n=513). A three-group latent class model was chosen due to minimal improvement in BIC criteria or clinical interpretability when compared to four-and five-group models and this was unchanged when the analysis was stratified by ethnicity. Maximum follow-up time in our study cohort was ten years, with median follow-up time at 2.9 years (IQR 1.0-5.4 years).

### Type 2 diabetes subgroups

Across the whole population, we characterised two T2DM subgroups identified in previous studies: ‘Mild Age-related Diabetes’ (MARD)’ was driven by age at diagnosis and was the most prevalent cluster (82% of the total population, n=26,294), and ‘Mild Obesity-related Diabetes MOD)’ was driven by BMI at onset (10%, n=3,059) (see Table 1 and Figure 1). We also identified a third subgroup, characterised by severe hyperglycaemia (determined by HbA1c) at diagnosis which we called ‘Severe Hyperglycaemic Diabetes (SHD)’ and was the least prevalent cluster (8%, n=2,578).

**Table 1.**
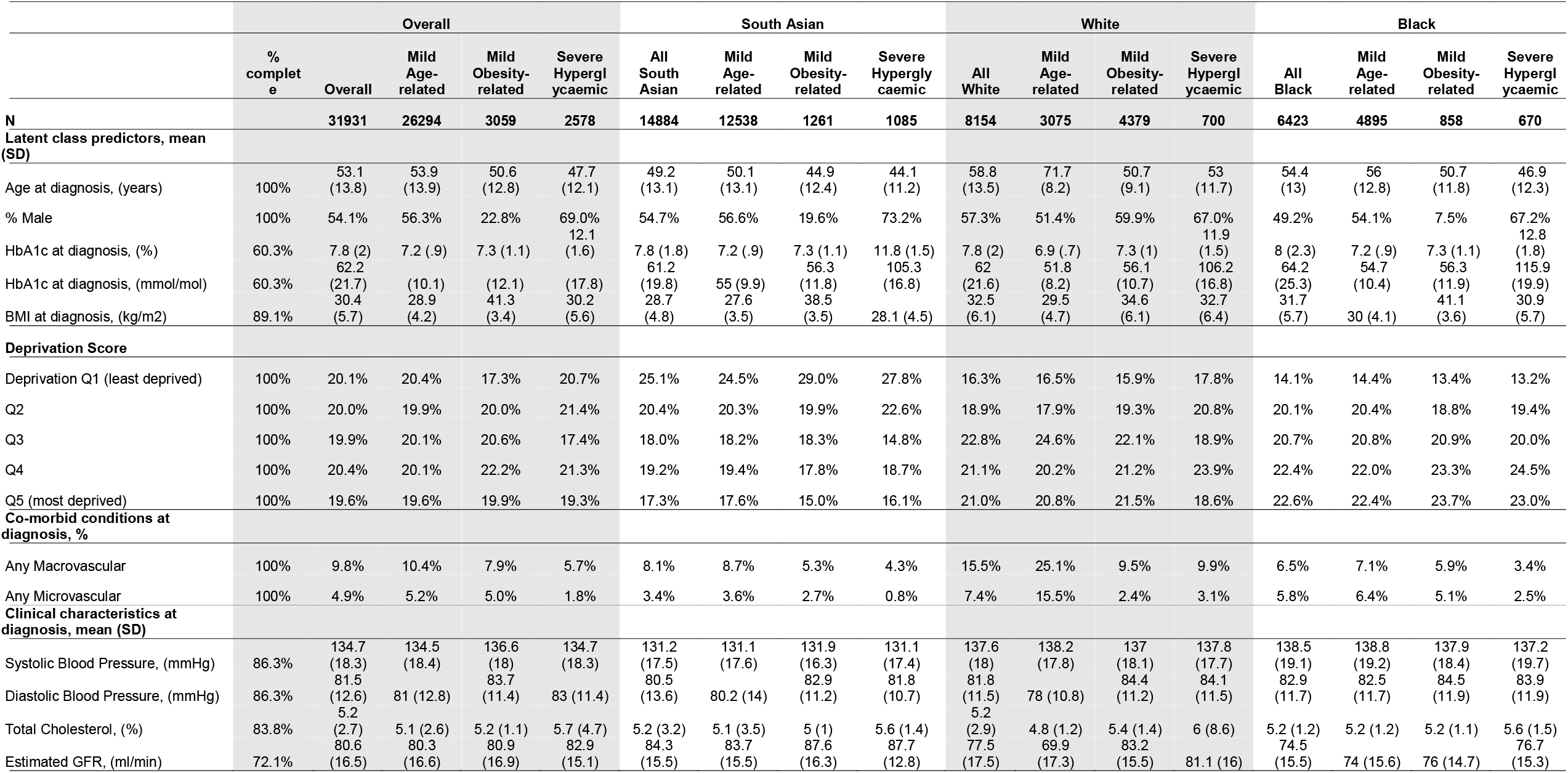
Baseline characteristics by T2DM subgroup for whole east London population and separately by ethnic group

**Figure 1:**
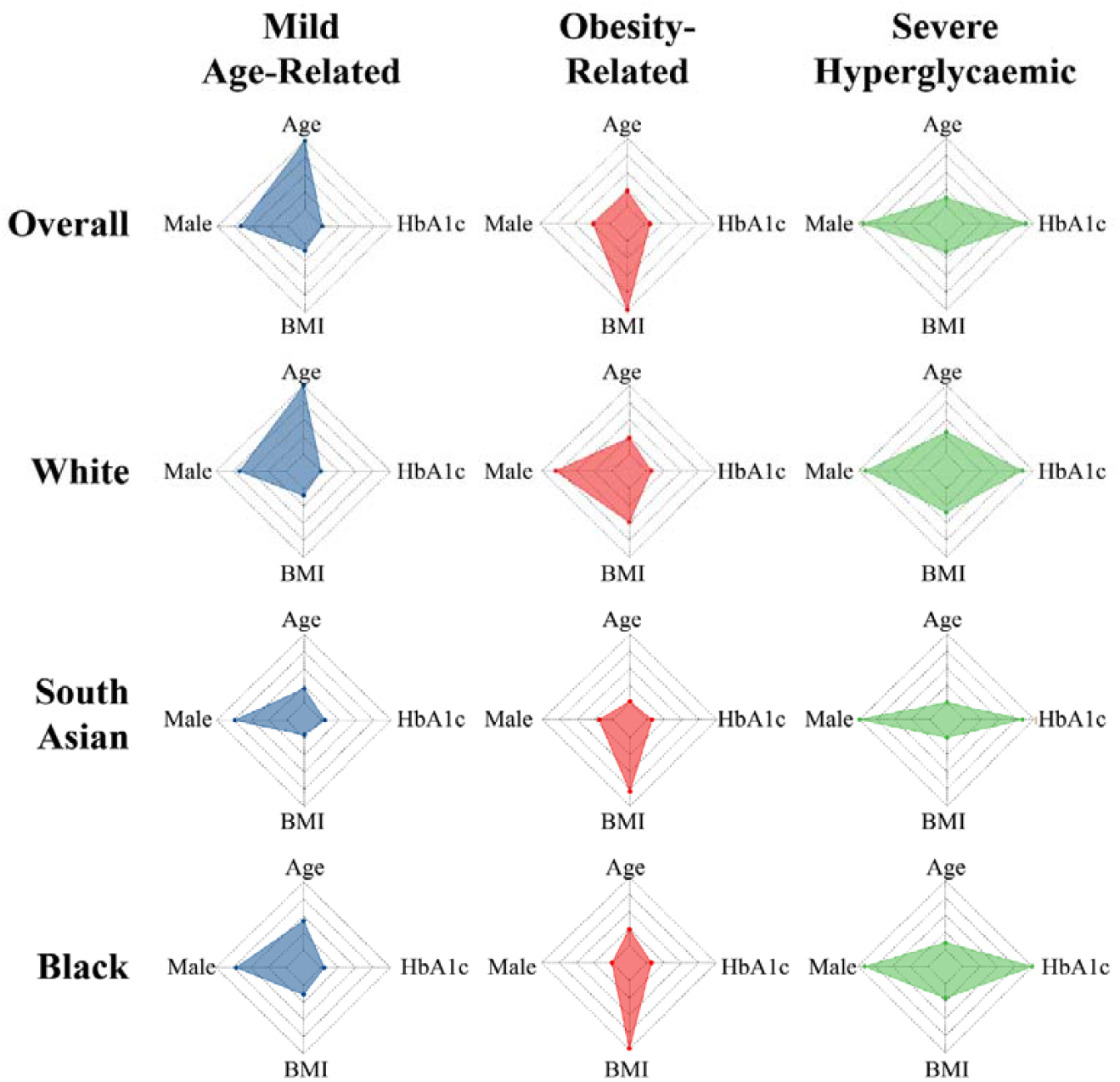
Radar plots to show distribution of age, body mass index (BMI), HbA1c, and sex (presented as proportion of males) across latent-class derived clusters in the overall cohort and by ethnicity. * The centre of each polygon represents the minimum value for that variable across all groups; the edge represents the maximum value. All polygons are plotted to the same scale. Overall cohort is 82.3% MARD, 9.6% MOD, 8.1% SHD; White ethnicity is 84.2% MARD, 8.5% MOD, 7.3% SHD; South Asian ethnicity is 37.7% MARD, 53.7% MOD, 8. SHD, Black ethnicity is 76.2% MARD, 13.4% MOD, 10.4% SHD.

### Ethnic differences in Type 2 diabetes subgroup membership

We then observed how cluster membership and clinical features varied by ethnicity (see Table 1 and Figure 1). In the white population, the MOD subgroup was the commonest (53% of individuals) and was characterized by severe obesity (mean BMI 34.6 kg/m^2^); 38% of the white population fell into the MARD subgroup (mean age 71.7 years), and 9% fell into the SHD subgroup (mean HbA1c 11.9%/106.2 mmol/mol). In contrast, amongst south Asians, the age-related subgroup was the commonest (84%) but was driven by a much younger mean age than in the White population (50.1 years). The MOD and SHD subgroups were uncommon (9% and 7%, respectively) in South Asians. As shown in Figure 1, the pattern and features of subgroups seen in South Asians were mirrored in the black African/Caribbean population, with 76% having MARD (mean age 56.0), 13% with MOD, and 10% with SHD. The clinical features driving MOD (BMI 41.4 kg/m2) and SHD (mean HbA1c 12.8%/115.9 mmol/mol) were more extreme in black African/Caribbean groups than in White and South Asian groups (Table 1.)

In both White and Black ethnic groups, the proportion of people with type 2 diabetes increased with increasing levels of deprivation. However, this gradient was reversed in the south Asian group with the majority of individuals contributing to the least deprived quintile. While the sex split was comparable between ethnic groups for the MARD and SHD subgroups, the MOD subgroup was 40% female for the White group, 80.4% female for the south Asian group and 92.5% female in the Black group. (Table 1)

### Time to vascular outcomes

Amongst those free from vascular disease at baseline, 4.3% developed macrovascular complications (n=1,094) and 28.5% developed microvascular complications during follow-up (Table S1). In comparison to the MARD subgroup, time to development of macrovascular disease was significantly increased in the MOD subgroup (HR 1.50, 95%CI 1.23-1.83), while time to development of microvascular disease was significantly increased in the SHD subgroup (HR 1.38, 95%CI 1.28-1.49) (Figure 2). These differences were most pronounced in the south Asian and Black groups, for whom subgroup differences in risk of microvascular and macrovascular outcomes were similar. For individuals of white ethnicity, no differences in the risk of either micro- or macrovascular disease by subgroup were evident. Despite the significant variation in age of onset across the MARD subgroups between the 3 ethnic categories, there was no differential association between MARD subgroup membership and vascular outcomes by ethnicity.

**Figure 2.**
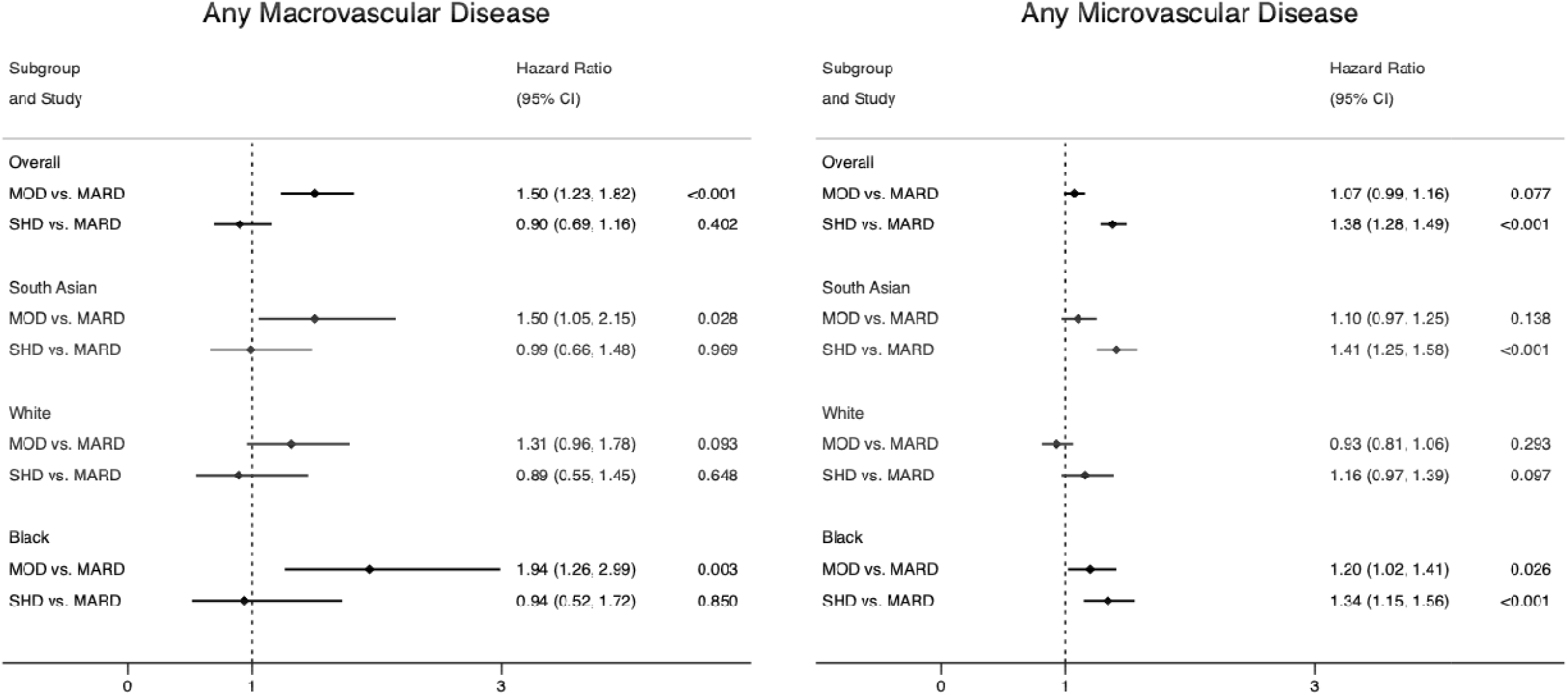
Risk of incident vascular complications by T2DM subgroup and ethnic group amongst those free of complications at baseline *All models age-sex adjusted. Macrovascular = any of CVD, stroke, heart failure, Microvascular = any of retinopathy, neuropathy, chronic kidney disease

### Time to treatment initiation

Amongst those free from treatment at baseline, 94% of those in the SHD subgroup, 82% of those in the MOD subgroup and 77% of those in the MARD subgroup initiated non-insulin antidiabetic treatment while 11.4% of those in the SHD subgroup, 6.5% of those in the MOD subgroup and 5.2% of those in the MARD subgroup-initiated insulin treatment. After adjustment for age and sex, initiation of non-insulin and insulin antidiabetic treatment was twice as fast in the SHD subgroup in comparison to the MARD subgroup (Non-insulin HR 1.92, 95%CI 1.83-2.02, Insulin HR 2.02, 95%CI 1.76-2.32), with differences between the MOD and MARD subgroups smaller in magnitude (Non-insulin HR 1.16, 95%CI 1.11-1.22, Insulin HR 1.02, 95%CI 0.87-1.21). Subgroup differences in age-sex adjusted time to treatment initiation were largest in the White population and smallest in the Black population (Table 2).

**Table 2.** Treatment initiation characteristics by diabetes subgroup and ethnicity

| Population | Non-Insulin Oral Antidiabetic Medications |  |  |  | Insulin |  |  |  |
| --- | --- | --- | --- | --- | --- | --- | --- | --- |
|  | Number eligible to initiate | Initiating treatment, (N, %) | Years to initiation, (mean, SD) | Relative risk vs. Mild-age related (age-sex adjusted) | Number eligible to initiate | Initiating treatment, (N, %) | Years to initiation, (mean, SD) | Relative risk vs. Mild-age related (age-sex adjusted) |
| Overall | 26,426 | 20,781 (78.6) | 0.9 (1.7) |  | 27,442 | 1,590 (5.8) | 3.7 (2.7) |  |
| Mild age-related | 21,860 | 16,781 (76.8) | 1.0 (1.7) | 1 (ref) | 22,623 | 1,170 (5.2) | 3.7 (2.7) | 1 (ref) |
| Mild Obesity-related | 2,455 | 2,016 (82.1) | 0.8 (1.6) | 1.16 (1.11-1.22), p <0.001 | 2,617 | 169 (6.5) | 3.8 (2.7) | 1.02 (0.87-1.21), p 0.778 |
| Severe Hyperglycaemic | 2,111 | 1,984 (94) | 0.3 (1.1) | 1.92 (1.83-2.02), p <0.001 | 2,202 | 251 (11.4) | 3.4 (2.6) | 2.02 (1.76-2.32), p <0.001 |
| South Asian | 11,921 | 9,832 (82.5) | 0.7 (1.4) |  | 12,445 | 563 (4.5) | 3.7 (2.6) |  |
| Mild age-related | 10,129 | 8,196 (80.9) | 0.8 (1.5) | 1 (ref) | 10,518 | 418 (4) | 3.7 (2.6) | 1 (ref) |
| Mild Obesity-related | 933 | 804 (86.2) | 0.6 (1.3) | 1.14 (1.06-1.23), p <0.001 | 1,017 | 60 (5.9) | 3.6 (2.5) | 0.98 (0.74-1.29), p 0.879 |
| Severe Hyperglycaemic | 859 | 832 (96.9) | 0.1 (.7) | 2.05 (1.90-2.20), p <0.001 | 910 | 85 (9.3) | 3.5 (2.5) | 2.12 (1.67-2.69), p <0.001 |
| White | 7,076 | 5,256 (74.3) | 1.1 (2) |  | 7,307 | 468 (6.4) | 3.9 (2.7) |  |
| Mild age-related | 2,734 | 1,714 (62.7) | 1.5 (2.2) | 1 (ref) | 2,837 | 111 (3.9) | 3.9 (2.7) | 1 (ref) |
| Mild Obesity-related | 3,749 | 3,000 (80) | 0.9 (1.7) | 1.40 (1.29-1.53), p <0.001 | 3,860 | 277 (7.2) | 3.9 (2.8) | 0.92 (0.68-1.25), p 0.597 |
| Severe Hyperglycaemic | 593 | 542 (91.4) | 0.4 (1.3) | 2.43 (2.17-2.72), p <0.001 | 610 | 80 (13.1) | 3.5 (2.6) | 2.00 (1.41-2.83), p <0.001 |
| Black | 5,400 | 4,152 (76.9) | 0.9 (1.7) |  | 5,558 | 448 (8.1) | 3.6 (2.7) |  |
| Mild age-related | 4,131 | 3,078 (74.5) | 1 (1.8) | 1 (ref) | 4,245 | 312 (7.3) | 3.6 (2.7) | 1 (ref) |
| Mild Obesity-related | 709 | 556 (78.4) | 0.8 (1.6) | 1.12 (1.02-1.23), p 0.022 | 739 | 49 (6.6) | 3.6 (2.7) | 0.72 (0.53-0.99), p 0.045 |
| Severe Hyperglycaemic | 560 | 518 (92.5) | 0.3 (1.2) | 1.81 (1.64-2.00), p <0.001 | 574 | 87 (15.2) | 3.3 (2.6) | 1.56 (1.21-2.00), p <0.001 |

\* The centre of each polygon represents the minimum value for that variable across all groups; the edge represents the maximum value. All polygons are plotted to the same scale. Overall cohort is 82.3% MARD, 9.6% MOD, 8.1% SHD; White ethnicity is 84.2% MARD, 8.5% MOD, 7.3% SHD; South Asian ethnicity is 37.7% MARD, 53.7% MOD, 8. SHD, Black ethnicity is 76.2% MARD, 13.4% MOD, 10.4% SHD.

### HbA1c trajectories over time

In the MARD and MOD subgroups, HbA1c was below 7.6%/60 mmol/mol at diagnosis and remained so over the first five years of follow-up. In the SHD subgroup, HbA1c was significantly elevated at diagnosis (>12.7%/90 mmol/mol) and brought down rapidly within the first 12 months, though never achieving target control. Patterns of HbA1c trajectories in each of the ethnic groups mirrored that of the overall population (Figure 3).

**Figure 3.**
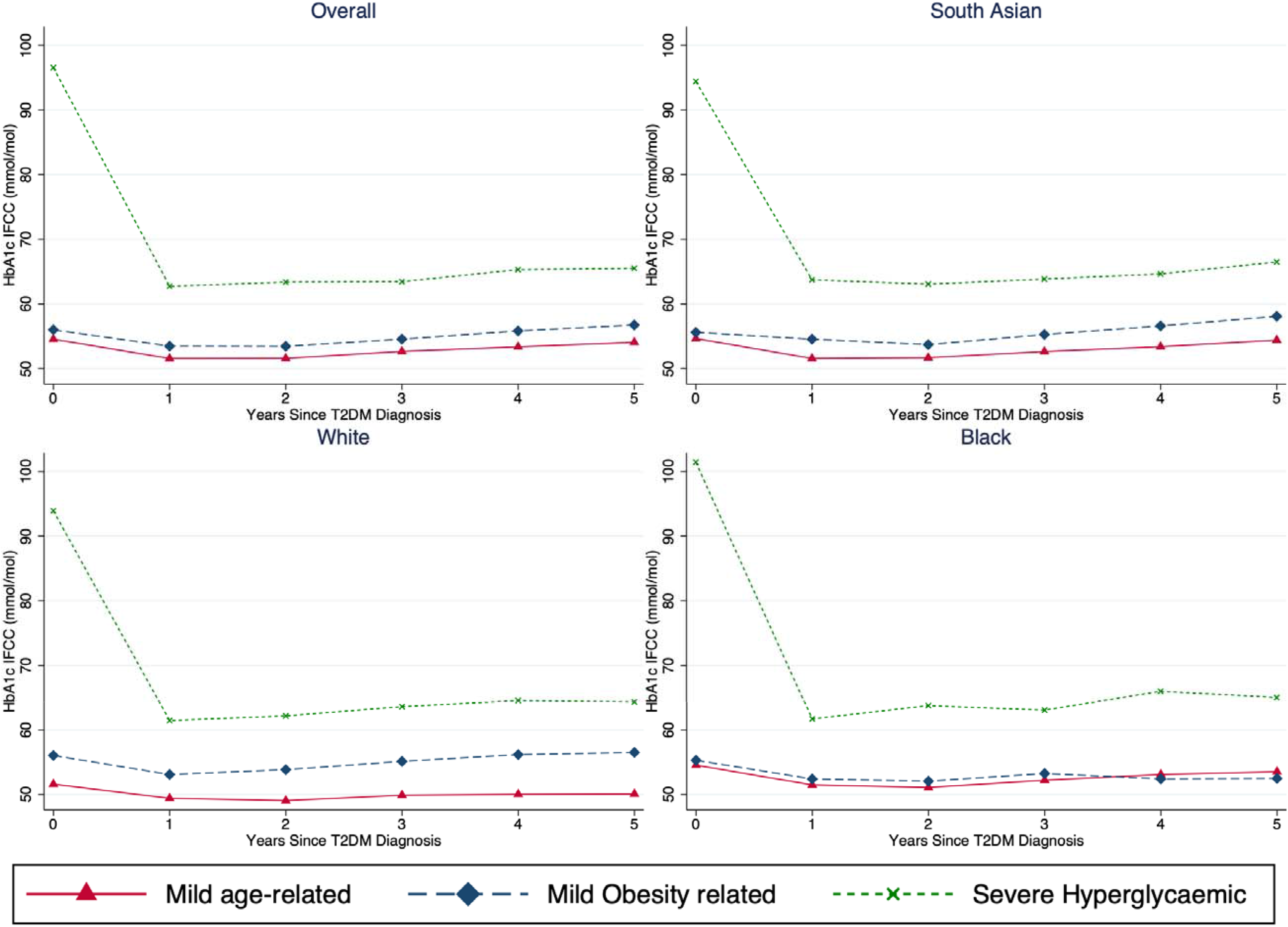
HbA1c trajectories by latent subgroup and ethnic group

## Discussion

In this observational cohort study, we identified three T2DM subgroups in an ethnically diverse UK population. Using only routinely recorded primary care clinical observations we replicated two subgroups previously reported in experimental and trial cohorts but were unable to identify subgroups based on insulin secretion or insulin resistance, as these rely on biomarkers not widely available in primary care settings.

We showed that T2DM subgroups defined at the time of diagnosis were strong predictors of clinically important differences in time to onset of vascular disease, time to initiation of antidiabetic treatment, and attainment of glycaemic control. People classified as having ‘Mild age-related’ diabetes at diagnosis had the slowest progression to vascular complications, slowest initiation of antidiabetic treatment, and best long-term glycaemic control. This association was observed consistently across all ethnic groups, despite age at diagnosis being 22 and 16 years earlier in south Asian and Black groups respectively than White Europeans. Future research will need to investigate the longer-term impact of South Asian and Black people in the MARD subgroup on vascular complications. People classified as having ‘Mild Obesity-related diabetes’ at diagnosis maintained target HbA1c control over the study duration, had faster treatment initiation than those in the MARD subgroup and had the fastest progression to macrovascular complications. Those classified as having ‘severe hyperglycaemic’ diabetes at diagnosis had persistently poor glycaemic control which never reached target thresholds and had the highest risk of microvascular outcomes despite having the fastest initiation of antidiabetic medication. Our findings suggest that people in the age-related subgroup may require less intensive clinical care processes, but that those in the severe hyperglycaemic group are likely to benefit from enhanced monitoring, support and management of their condition.

There were significant differences in the clinical features of diabetes subgroups according to ethnicity. For example, the MOD subgroup had disproportionately more women from south Asian and Black ethnicities than Whites. The age at diagnosis of people with MARD was 20 years younger in south Asian and Black ethnicity than Whites. Our study indicates that south Asians and Blacks in the MOD and SHD subgroups do have a stronger association with macrovascular and microvascular complications, respectively, than Whites. Future research in larger multi-ethnic populations will be needed to further investigate the impact of sex and age of diabetes onset on disease outcomes.

### Comparison with existing literature

Previous studies have identified four type 2 diabetes subgroups: Severe insulin deficient diabetes (SIDD), severe insulin resistant diabetes (SIRD), mild obesity-related diabetes (MOD), and mild age-related diabetes (MARD). While these studies have generated reproducible findings, they are limited to experimental populations of largely White Europeans. In the UK the clinical measures necessary to replicate these subgroups are only available in secondary care data, limiting their usefulness for diabetes management which largely takes place in primary care settings.

Using routinely collected data, we identified subgroups which closely align with those previously reported. Our ‘Mild Age-Related Diabetes’ and ‘Mild Obesity-Related Diabetes’ subgroups resembled the MARD and MOD clusters. Our ‘Severe Hyperglycaemic Diabetes’ cluster was specific to our study and is likely to include the previously-reported ‘Severe Insulin Deficient Diabetes’ and ‘Severe Insulin Resistant Diabetes’ clusters.

### Strengths

Our study benefits from a significantly larger and more ethnically diverse population than many earlier studies used for identifying T2DM subgroups.^3,5,6^ Furthermore, ethnicity was recorded for 98% of the study population, ensuring that the ethnicity specific T2DM analyses were unlikely to be biased. This study captured all people with type 2 diabetes registered within a contiguous geographic area representative of the general population and other urban centres in the UK which are also ethnically and socially diverse. Furthermore, all practices contributing to our study were following standard diabetes management guidelines as outlined by the National Institute for Health and Care Excellence^14^ and performance standards as per the Quality and Outcomes Framework.^15^

### Limitations

We did not include measures of serum high-density lipoprotein and triglycerides in our cluster analysis as these are not uniformly collected in routine primary care practice. Their inclusion would have added further refinement of diabetes subgroups as they are surrogate measures of insulin resistance. The east London Primary Care Database is subject to the same strengths and biases as all routine data.^16^ It is possible that by using diagnostic codes to define diabetes, some individuals with type 1 diabetes may have been misclassified as having type 2 diabetes. We identified prescriptions for antidiabetic medications issued in primary care but were unable to determine whether prescriptions were filled or taken as indicated. Finally, we did not have access to linked secondary care data and may have missed acute vascular events coded in hospital settings only.

### Conclusions

We have demonstrated that pragmatic Type 2 diabetes subgroups can be generated using real-world primary care data and can identify important differences in clinical characteristics and vascular outcomes. Our findings have wider generalisability to national and global populations. The identification of these subgroups provides a useful heuristic for characterising differences between patients at population level, including in ethnically diverse populations. Identification of these subgroups at diagnosis and could help move away from a “one size fits all” care pathway and instead offer stratified care pathway that is readily enabled by clinical data systems. This stratification could enhance the care of those most at risk of complications, and de-intensify care for those who are not. Opportunities to stratify care are particularly relevant in the context of healthcare services constrained in time and financial resources in which many people with T2DM and clinicians managing their care feel their care needs are not met.^17^ The ability to apply data-driven clustering to real-world data offers wider generalisability to other chronic diseases largely managed in primary care such as hypertension and chronic kidney disease.

Important next steps are to reproduce these findings in other multi-ethnic populations, using larger sample sizes, longer follow-up duration and lipid profile measures to reproduce these findings at scale. Subsequently, empirical evaluation of subgroup-stratified care using a cluster randomised controlled trial with long-term measurement of outcomes is likely to be necessary.

## Supporting information

(see supplementary material)

## Data Availability

The data used for this study comprises anonymised clinical records derived from the east London Database. Only the authors have access to the study data. Code lists are available in the supplementary materials. Researchers should contact the Clinical Effectiveness Group at QMUL to request access to data.

## Author contributions

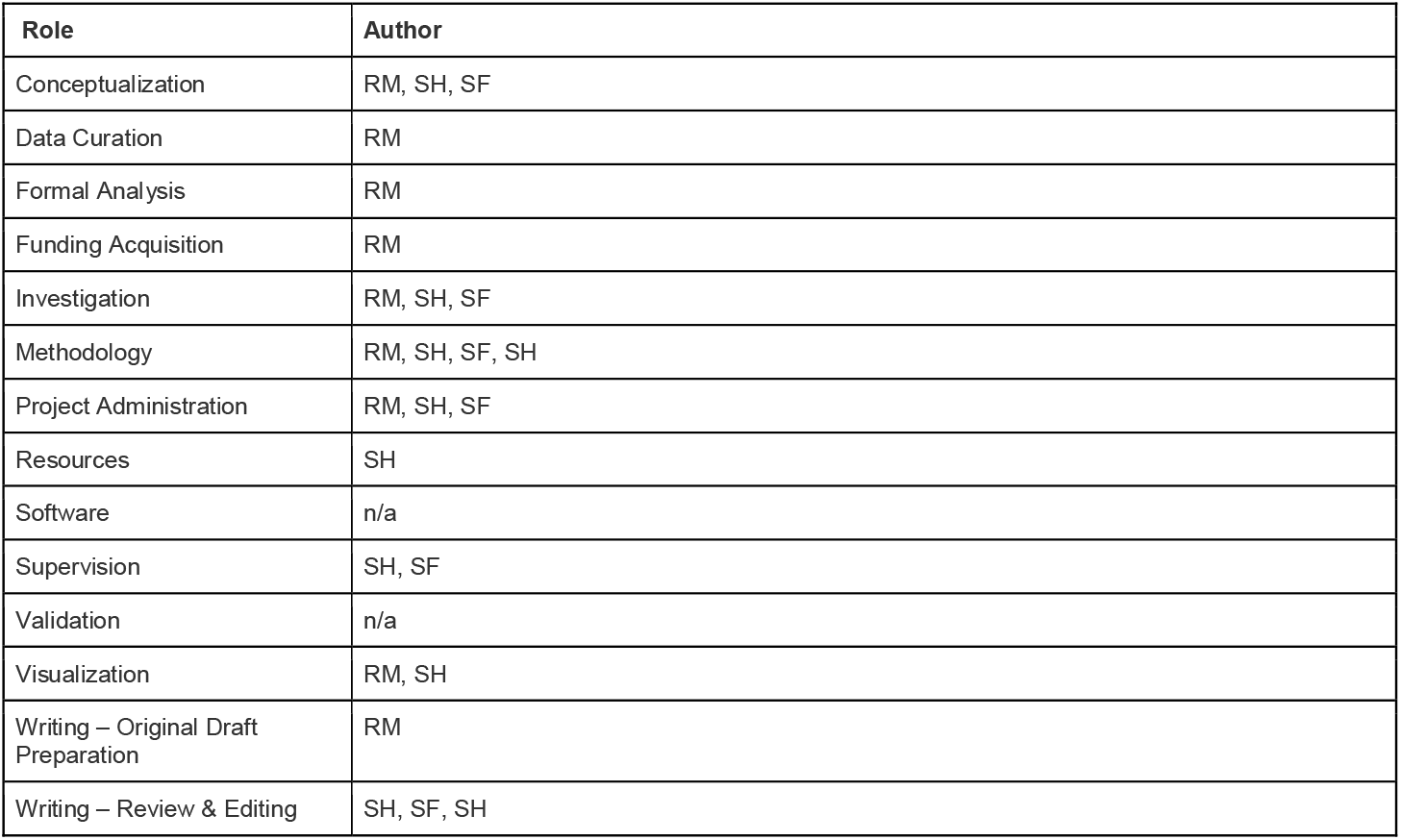

