## Supplementary material for "Characterisation of type 2 diabetes subgroups and their association with ethnicity and clinical outcomes: a UK real-world data study using the East London Database": (see supplementary material)

### Supplementary Table S1: Time to onset of vascular complications

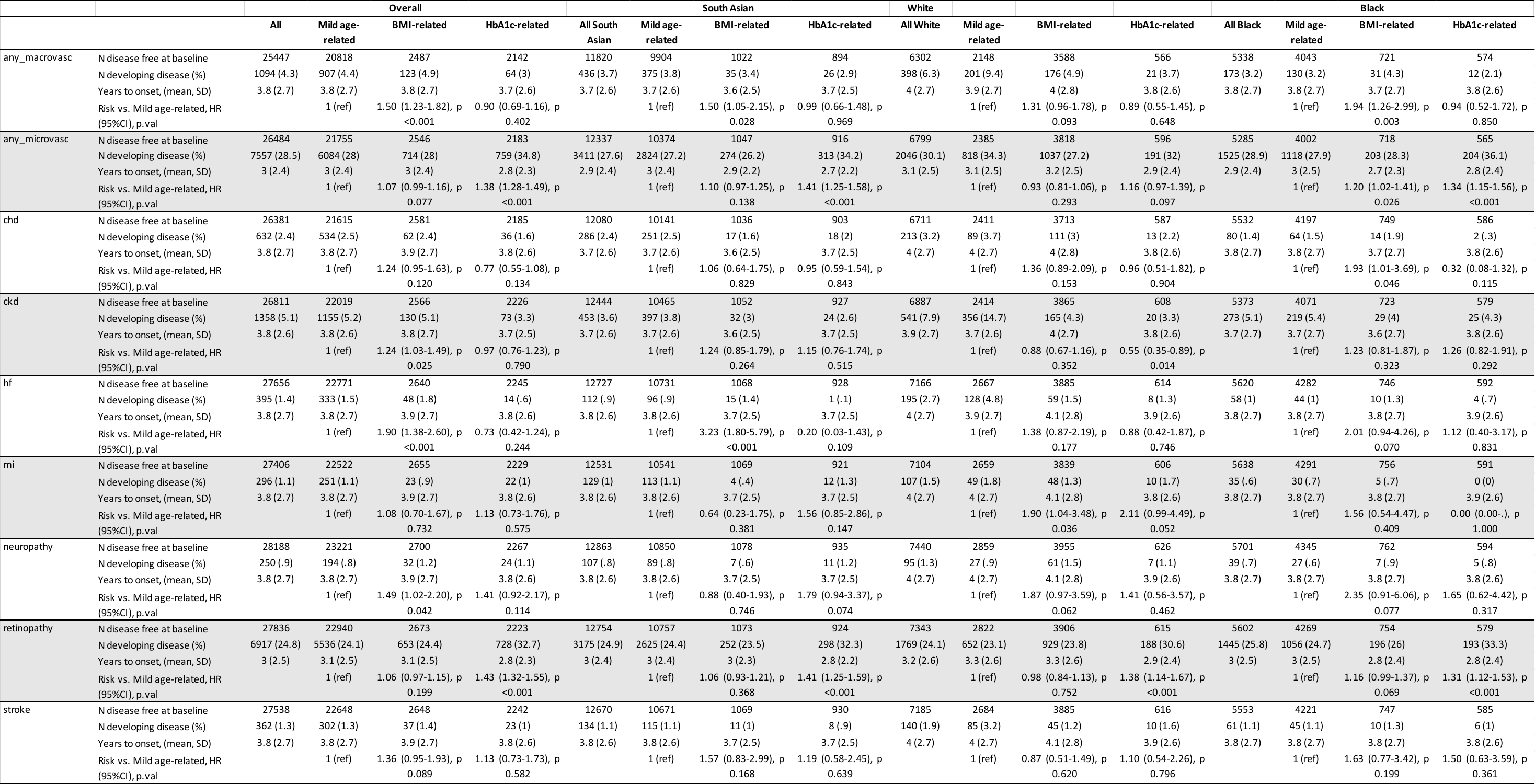

### Supplementary Table S2: Codelists

#### Demographic Characteristics

| Variable | Read code |
| --- | --- |
| Gender | Registration Data |
| Age | Registration Data |
| Ethnicity | 9S%, 9i% |
| IMD2010 | Registration Data |
| First registration with GP | Registration Data |
| Last registration with GP | Registration Data |
| Death date | Registration Data |

#### Laboratory results

| BMI value and date | 22K% |
| --- | --- |
| HbA1c | 42W5 (IFFC), 42W4 (DCCT), |
| Cholesterol | 44P |
| Systolic Blood Pressure | 2469 |
| Diastolic Blood Pressure | 246A |
| eGFR | 451G 451K |

#### Diagnostic Codes

| Ischaemic Heart Disease\CHD | G3, G30%, G31%, G32, G33%, G34%, G35%, G38%, G39%, G3y%, G3z, Gyu3% |
| --- | --- |
| Retinopathy | 2BB% F4213 |
| Neuropathy | F367. F4H34 F37.. F170. F36.. |

##### Type 2 Diabetes Mellitus

| Read code | Read term |
| --- | --- |
| C10FA | Type 2 diabetes mellitus with mononeuropathy |
| C10F1 | Type 2 diabetes mellitus with ophthalmic complications |
| C10F3 | Type 2 diabetes mellitus with multiple complications |
| C10FG | Type II diabetes mellitus with arthropathy |
| C10FM | Type 2 diabetes mellitus with persistent microalbuminuria |
| C10F5 | Type II diabetes mellitus with gangrene |
| C10F0 | Type 2 diabetes mellitus with renal complications |
| C10F3 | Type II diabetes mellitus with multiple complications |
| C10F5 | Type 2 diabetes mellitus with gangrene |
| C10FA | Type II diabetes mellitus with mononeuropathy |
| C10FC | Type II diabetes mellitus with nephropathy |
| C10FK | Hyperosmolar non-ketotic state in type 2 diabetes mellitus |
| C10FP | Type 2 diabetes mellitus with ketoacidotic coma |
| C10F2 | Type 2 diabetes mellitus with neurological complications |
| C10F2 | Type II diabetes mellitus with neurological complications |
| C10FB | Type 2 diabetes mellitus with polyneuropathy |
| C10FH | Type 2 diabetes mellitus with neuropathic arthropathy |
| C10FB | Type II diabetes mellitus with polyneuropathy |
| C10F. | Type II diabetes mellitus |
| C10F6 | Type 2 diabetes mellitus with retinopathy |
| C10FF | Type II diabetes mellitus with peripheral angiopathy |
| C10FM | Type II diabetes mellitus with persistent microalbuminuria |
| C10F9 | Type 2 diabetes mellitus without complication |
| C10FL | Type 2 diabetes mellitus with persistent proteinuria |
| C10F4 | Type 2 diabetes mellitus with ulcer |
| C10F9 | Type II diabetes mellitus without complication |
| C10FE | Type 2 diabetes mellitus with diabetic cataract |
| C10FN | Type 2 diabetes mellitus with ketoacidosis |
| C10F6 | Type II diabetes mellitus with retinopathy |
| C10F0 | Type II diabetes mellitus with renal complications |
| C10FG | Type 2 diabetes mellitus with arthropathy |
| C10FC | Type 2 diabetes mellitus with nephropathy |
| C10FD | Type II diabetes mellitus with hypoglycaemic coma |
| C10FE | Type II diabetes mellitus with diabetic cataract |
| C10F1 | Type II diabetes mellitus with ophthalmic complications |
| C10FJ | Insulin treated Type II diabetes mellitus |
| C10F4 | Type II diabetes mellitus with ulcer |
| C10FQ | Type 2 diabetes mellitus with exudative maculopathy |
| C10FR | Type 2 diabetes mellitus with gastroparesis |
| C10FF | Type 2 diabetes mellitus with peripheral angiopathy |
| C10FJ | Insulin treated Type 2 diabetes mellitus |
| C10F. | Type 2 diabetes mellitus |
| C10FD | Type 2 diabetes mellitus with hypoglycaemic coma |
| C10F7 | Type II diabetes mellitus - poor control |
| C10FL | Type II diabetes mellitus with persistent proteinuria |
| C10F7 | Type 2 diabetes mellitus - poor control |
| C110. | Self-induced hyperinsulinism |
| C1070 | Diabetes mellitus, juvenile +peripheral circulatory disorder |
| C112. | Hypoglycaemia unspecified |
| C1001 | Diabetes mellitus, adult onset, no mention of complication |
| C1071 | Diabetes mellitus, adult, + peripheral circulatory disorder |
| C1031 | Diabetes mellitus, adult onset, with ketoacidotic coma |
| L180X | Pre-existing diabetes mellitus, unspecified |
| C1051 | Diabetes mellitus, adult onset, + ophthalmic manifestation |
| C1001 | Non-insulin dependent diabetes mellitus |
| C112. | Reactive hypoglycaemia NOS |
| C112. | Hypoglycaemia unspecified NOS |
| C1001 | Maturity onset diabetes |
| C1061 | Diabetes mellitus, adult onset, + neurological manifestation |
| C112. | Spontaneous hypoglycaemia NOS |
| C1072 | Diabetes mellitus, adult with gangrene |
| C1021 | Diabetes mellitus, adult onset, with hyperosmolar coma |
| C1041 | Diabetes mellitus, adult onset, with renal manifestation |
| C109K | Hyperosmolar non-ketotic state in type 2 diabetes mellitus |
| C109J | Insulin treated non-insulin dependent diabetes mellitus |
| C109B | Type II diabetes mellitus with polyneuropathy |
| C109G | Non-insulin dependent diabetes mellitus with arthropathy |
| C109A | Non-insulin dependent diabetes mellitus with mononeuropathy |
| C109. | NIDDM - Non-insulin dependent diabetes mellitus |
| C1090 | Type II diabetes mellitus with renal complications |
| C109C | Type 2 diabetes mellitus with nephropathy |
| C109F | Non-insulin-dependent d m with peripheral angiopath |
| C1091 | Type 2 diabetes mellitus with ophthalmic complications |
| C1094 | Non-insulin dependent diabetes mellitus with ulcer |
| C109E | Type 2 diabetes mellitus with diabetic cataract |
| C1091 | Type II diabetes mellitus with ophthalmic complications |
| C109J | Insulin treated Type II diabetes mellitus |
| C109A | Type II diabetes mellitus with mononeuropathy |
| C1096 | Type 2 diabetes mellitus with retinopathy |
| C1090 | Non-insulin-dependent diabetes mellitus with renal comps |
| C1092 | Type II diabetes mellitus with neurological complications |
| C109G | Type II diabetes mellitus with arthropathy |
| C1097 | Non-insulin dependent diabetes mellitus - poor control |
| L1806 | Pre-existing diabetes mellitus, non-insulin-dependent |
| C109H | Type II diabetes mellitus with neuropathic arthropathy |
| C1093 | Non-insulin-dependent diabetes mellitus with multiple comps |
| C109E | Non-insulin depend diabetes mellitus with diabetic cataract |
| C1074 | NIDDM with peripheral circulatory disorder |
| C1092 | Non-insulin-dependent diabetes mellitus with neuro comps |
| C109. | Type 2 diabetes mellitus |
| C109B | Non-insulin dependent diabetes mellitus with polyneuropathy |
| C1090 | Type 2 diabetes mellitus with renal complications |
| C109D | Non-insulin dependent diabetes mellitus with hypoglyca coma |
| C109F | Type 2 diabetes mellitus with peripheral angiopathy |
| C109C | Non-insulin dependent diabetes mellitus with nephropathy |
| C1095 | Non-insulin dependent diabetes mellitus with gangrene |
| C1097 | Type II diabetes mellitus - poor control |
| C109G | Type 2 diabetes mellitus with arthropathy |
| C1096 | Non-insulin-dependent diabetes mellitus with retinopathy |
| C1095 | Type 2 diabetes mellitus with gangrene |
| C109D | Type II diabetes mellitus with hypoglycaemic coma |
| C10y1 | Diabetes mellitus, adult, + other specified manifestation |
| C1094 | Type II diabetes mellitus with ulcer |
| C109H | Non-insulin dependent d m with neuropathic arthropathy |
| C109J | Insulin treated Type 2 diabetes mellitus |
| C109E | Type II diabetes mellitus with diabetic cataract |
| C1094 | Type 2 diabetes mellitus with ulcer |
| C1091 | Non-insulin-dependent diabetes mellitus with ophthalm comps |
| C109D | Type 2 diabetes mellitus with hypoglycaemic coma |
| C109F | Type II diabetes mellitus with peripheral angiopathy |
| C109C | Type II diabetes mellitus with nephropathy |
| C1097 | Type 2 diabetes mellitus - poor control |
| C109. | Type II diabetes mellitus |
| C109. | Non-insulin dependent diabetes mellitus |
| C1099 | Non-insulin-dependent diabetes mellitus without complication |
| C10z1 | Diabetes mellitus, adult onset, + unspecified complication |
| C1095 | Type II diabetes mellitus with gangrene |
| C1092 | Type 2 diabetes mellitus with neurological complications |
| C1096 | Type II diabetes mellitus with retinopathy |
| C109H | Type 2 diabetes mellitus with neuropathic arthropathy |

##### Myocardial Infarction

| Read code | Read term |
| --- | --- |
| 14AP.00 | History of ventricular tachycardia |
| 14AD.00 | H/O ventricular fibrillation |
| 7937600 | Removal of intermal cardiac defibrillator |
| 793F200 | Resiting of lead of cardioverter defibrillator |
| 793F300 | Renewal of cardioverter defibrillator |
| 793F400 | Removal of cardioverter defibrillator |
| 3282 | ECG: ventricular tachycardia |
| G571.11 | Ventricular tachycardia |
| 3283 | ECG: ventricular fibrillation |
| 7L1H.13 | Defibrillation |
| G574.00 | Ventricular fibrillation and flutter |
| G574000 | Ventricular fibrillation |
| G574011 | Cardiac arrest-ventricular fibrillation |
| G574z00 | Ventricular fibrillation and flutter NOS |
| 7937500 | Implantation of intermal cardiac defibrillator |
| 793F.00 | Cardioverter defibrillator introduced through the vein |
| 793F000 | Implantat cardioverter defibrillator us one electrode lead |
| 793F100 | Implant cardioverter defibrillator using two electrode leads |
| 793F500 | Implantat cardiovert defibrillator us three electrode leads |
| 793Fy00 | Other specified cardioverter defibrillator intro thr vein |
| 793Fz00 | Cardioverter defibrillator introduced through the vein NOS |
| 2241 | O/E - collapse -cardiac arrest |
| 7932111 | Cardiac massage - open |
| 7L1H600 | Advanced cardiopulmonary resuscitation |
| 853..00 | Cardiac massage - extermal |
| 8531 | Closed cardiac massage alone |
| 8532 | Closed cardiac massage+ventil. |
| 8532.11 | Cardiopulmonary resuscitation |
| 853Z.00 | Extermal cardiac massage NOS |
| G575.00 | Cardiac arrest |
| G575.11 | Cardio-respiratory arrest |
| G575.12 | Asystole |
| G575000 | Cardiac arrest with successful resuscitation |
| G575200 | Electromechanical dissociation with successful resuscitation |
| G575300 | Electromechanical dissociation |
| G575z00 | Cardiac arrest, unspecified |
| SP11000 | Cardiac arrest as a complication of care |
| 889A.00 | Diab mellit insulin-glucose infus acute myocardial infarct |
| G30..00 | Acute myocardial infarction |
| G30..11 | Attack - heart |
| G30..12 | Coronary thrombosis |
| G30..13 | Cardiac rupture following myocardial infarction (MI) |
| G30..14 | Heart attack |
| G30..15 | MI - acute myocardial infarction |
| G30..16 | Thrombosis - coronary |
| G30..17 | Silent myocardial infarction |
| G300.00 | Acute anterolateral infarction |
| G301.00 | Other specified anterior myocardial infarction |
| G301000 | Acute anteroapical infarction |
| G301100 | Acute anteroseptal infarction |
| G301z00 | Anterior myocardial infarction NOS |
| G302.00 | Acute inferolateral infarction |
| G303.00 | Acute inferoposterior infarction |
| G304.00 | Posterior myocardial infarction NOS |
| G305.00 | Lateral myocardial infarction NOS |
| G306.00 | True posterior myocardial infarction |
| G307.00 | Acute subendocardial infarction |
| G307000 | Acute non-Q wave infarction |
| G308.00 | Inferior myocardial infarction NOS |
| G309.00 | Acute Q-wave infarct |
| G30A.00 | Mural thrombosis |
| G30B.00 | Acute posterolateral myocardial infarction |
| G30X.00 | Acute transmural myocardial infarction of unspecif site |
| G30y.00 | Other acute myocardial infarction |
| G30y000 | Acute atrial infarction |
| G30y100 | Acute papillary muscle infarction |
| G30y200 | Acute septal infarction |
| G30yz00 | Other acute myocardial infarction NOS |
| G30z.00 | Acute myocardial infarction NOS |
| G310.11 | Dressler's syndrome |
| G31y100 | Microinfarction of heart |
| G35..00 | Subsequent myocardial infarction |
| G350.00 | Subsequent myocardial infarction of anterior wall |
| G351.00 | Subsequent myocardial infarction of inferior wall |
| G353.00 | Subsequent myocardial infarction of other sites |
| G35X.00 | Subsequent myocardial infarction of unspecified site |
| G36..00 | Certain current complication follow acute myocardial infarct |
| G360.00 | Haemopericardium/current comp folow acut myocard infarct |
| G361.00 | Atrial septal defect/curr comp folow acut myocardal infarct |
| G362.00 | Ventric septal defect/curr comp fol acut myocardal infarctn |
| G363.00 | Ruptur cardiac wall w'out haemopericard/cur comp fol ac MI |
| G364.00 | Ruptur chordae tendinae/curr comp fol acute myocard infarct |
| G365.00 | Rupture papillary muscle/curr comp fol acute myocard infarct |
| G366.00 | Thrombosis atrium,auric append&vent/curr comp foll acute MI |
| G38..00 | Postoperative myocardial infarction |
| G380.00 | Postoperative transmural myocardial infarction anterior wall |
| G381.00 | Postoperative transmural myocardial infarction inferior wall |
| G384.00 | Postoperative subendocardial myocardial infarction |
| G38z.00 | Postoperative myocardial infarction, unspecified |
| G501.00 | Post infarction pericarditis |
| Gyu3400 | [X]Acute transmural myocardial infarction of unspecif site |
| G307100 | Acute non-ST segment elevation myocardial infarction |
| G30X000 | Acute ST segment elevation myocardial infarction |

##### Stroke

| Read code | Read term |
| --- | --- |
| 662o.00 | Haemorrhagic stroke monitoring |
| G681.00 | Sequelae of intracerebral haemorrhage |
| G682.00 | Sequelae of other nontraumatic intracranial haemorrhage |
| G61..00 | Intracerebral haemorrhage |
| G61..11 | CVA - cerebrovascular accid due to intracerebral haemorrhage |
| G61..12 | Stroke due to intracerebral haemorrhage |
| G610.00 | Cortical haemorrhage |
| G611.00 | Intermal capsule haemorrhage |
| G612.00 | Basal nucleus haemorrhage |
| G613.00 | Cerebellar haemorrhage |
| G614.00 | Pontine haemorrhage |
| G616.00 | Extermal capsule haemorrhage |
| G617.00 | Intracerebral haemorrhage, intraventricular |
| G618.00 | Intracerebral haemorrhage, multiple localized |
| G61X.00 | Intracerebral haemorrhage in hemisphere, unspecified |
| G61X000 | Left sided intracerebral haemorrhage, unspecified |
| G61X100 | Right sided intracerebral haemorrhage, unspecified |
| G61z.00 | Intracerebral haemorrhage NOS |
| Gyu6200 | [X]Other intracerebral haemorrhage |
| Gyu6F00 | [X]Intracerebral haemorrhage in hemisphere, unspecified |
| G601.00 | Subarachnoid haemorrhage from carotid siphon and bifurcation |
| G602.00 | Subarachnoid haemorrhage from middle cerebral artery |
| G60X.00 | Subarachnoid haemorrh from intracranial artery, unspecif |
| 7017000 | Evacuation of subdural haematoma |
| G621.00 | Subdural haemorrhage - nontraumatic |
| G622.00 | Subdural haematoma - nontraumatic |
| G623.00 | Subdural haemorrhage NOS |
| S62..13 | Subdural haemorrhage following injury |
| S622.00 | Closed traumatic subdural haemorrhage |
| S623.00 | Open traumatic subdural haemorrhage |
| S628.00 | Traumatic subdural haemorrhage |
| S629.00 | Traumatic subdural haematoma |
| S629000 | Traumatic subdural haematoma without open intracranial wound |
| S629100 | Traumatic subdural haematoma with open intracranial wound |
| 7032000 | Evacuation of extradural haematoma |
| G620.00 | Extradural haemorrhage - nontraumatic |
| S62..11 | Extradural haemorrhage following injury |
| S624.00 | Closed traumatic extradural haemorrhage |
| S624.11 | Epidural haematoma following injury |
| S625.00 | Open traumatic extradural haemorrhage |
| S626.00 | Epidural haemorrhage |
| S62A.00 | Traumatic extradural haematoma |
| G62..00 | Other and unspecified intracranial haemorrhage |
| G62z.00 | Intracranial haemorrhage NOS |
| A94y600 | Rupture of syphilitic cerebral aneurysm |
| S62..00 | Cerebral haemorrhage following injury |
| S62..14 | Traumatic cerebral haemorrhage |
| S62z.00 | Cerebral haemorrhage following injury NOS |
| S63..00 | Other cerebral haemorrhage following injury |
| S63z.00 | Other cerebral haemorrhage following injury NOS |
| 1477 | H/O: cerebrovascular disease |
| 1JA1.00 | Suspected cerebrovascular disease |
| 7N44000 | [SO]Carotid artery |
| 7N44500 | [SO]Common carotid artery |
| 7N44600 | [SO]Intermal carotid artery |
| 7N44L00 | [SO]Carotid artery NEC |
| 7N48200 | [SO]Carotid body |
| 7NB6400 | [SO]Extermal carotid artery |
| 7NB6500 | [SO]Common carotid artery |
| 7NB6600 | [SO]Intermal carotid artery |
| 7NBC.00 | [SO]Branch of extermal carotid artery |
| F14..11 | Cerebellar disease |
| G65..13 | Vertebro-basilar insufficiency |
| G650.00 | Basilar artery syndrome |
| G650.11 | Insufficiency - basilar artery |
| G651.00 | Vertebral artery syndrome |
| G651000 | Vertebro-basilar artery syndrome |
| G653.00 | Carotid artery syndrome hemispheric |
| G654.00 | Multiple and bilateral precerebral artery syndromes |
| G656.00 | Vertebrobasilar insufficiency |
| G673200 | Carotid artery dissection |
| G72y000 | Aneurysm of common carotid art |
| G72y100 | Aneurysm of extermal carotid artery |
| G72y200 | Aneurysm of intermal carotid artery |
| G769.00 | Anterior spinal and vertebral artery compression syndromes |
| N1...00 | Vertebral column syndromes |
| N111.11 | Vertebral artery compression syndrome |
| E004.00 | Arteriosclerotic dementia |
| E004000 | Uncomplicated arteriosclerotic dementia |
| E004100 | Arteriosclerotic dementia with delirium |
| E004200 | Arteriosclerotic dementia with paranoia |
| E004300 | Arteriosclerotic dementia with depression |
| E004z00 | Arteriosclerotic dementia NOS |
| Eu01.11 | [X]Arteriosclerotic dementia |
| G6...00 | Cerebrovascular disease |
| G63..00 | Precerebral arterial occlusion |
| G63..11 | Infarction - precerebral |
| G63..12 | Stenosis of precerebral arteries |
| G630.00 | Basilar artery occlusion |
| G631.00 | Carotid artery occlusion |
| G631.11 | Stenosis, carotid artery |
| G631.12 | Thrombosis, carotid artery |
| G632.00 | Vertebral artery occlusion |
| G633.00 | Multiple and bilateral precerebral arterial occlusion |
| G634.00 | Carotid artery stenosis |
| G63y.00 | Other precerebral artery occlusion |
| G63y000 | Cerebral infarct due to thrombosis of precerebral arteries |
| G63y100 | Cerebral infarction due to embolism of precerebral arteries |
| G63z.00 | Precerebral artery occlusion NOS |
| G64..00 | Cerebral arterial occlusion |
| G64..12 | Infarction - cerebral |
| G640.00 | Cerebral thrombosis |
| G640000 | Cerebral infarction due to thrombosis of cerebral arteries |
| G641.00 | Cerebral embolism |
| G641.11 | Cerebral embolus |
| G641000 | Cerebral infarction due to embolism of cerebral arteries |
| G64z.00 | Cerebral infarction NOS |
| G64z.11 | Brainstem infarction NOS |
| G64z.12 | Cerebellar infarction |
| G64z000 | Brainstem infarction |
| G64z200 | Left sided cerebral infarction |
| G64z300 | Right sided cerebral infarction |
| G64z400 | Infarction of basal ganglia |
| G660.00 | Middle cerebral artery syndrome |
| G661.00 | Anterior cerebral artery syndrome |
| G662.00 | Posterior cerebral artery syndrome |
| G67..00 | Other cerebrovascular disease |
| G671100 | Chronic cerebral ischaemia |
| G671z00 | Generalised ischaemic cerebrovascular disease NOS |
| G676000 | Cereb infarct due cerebral venous thrombosis, nonpyogenic |
| G677.00 | Occlusion/stenosis cerebral arts not result cerebral infarct |
| G677000 | Occlusion and stenosis of middle cerebral artery |
| G677100 | Occlusion and stenosis of anterior cerebral artery |
| G677200 | Occlusion and stenosis of posterior cerebral artery |
| G677300 | Occlusion and stenosis of cerebellar arteries |
| G677400 | Occlusion+stenosis of multiple and bilat cerebral arteries |
| G678.00 | Cereb autosom dominant arteriop subcort infarcts leukoenceph |
| G679.00 | Small vessel cerebrovascular disease |
| G67y.00 | Other cerebrovascular disease OS |
| G67z.00 | Other cerebrovascular disease NOS |
| G68..00 | Late effects of cerebrovascular disease |
| G68W.00 | Sequelae/other + unspecified cerebrovascular diseases |
| G6y..00 | Other specified cerebrovascular disease |
| G6z..00 | Cerebrovascular disease NOS |
| G70y000 | Carotid artery atherosclerosis |
| G70y011 | Carotid artery disease |
| Gyu6.00 | [X]Cerebrovascular diseases |
| G683.00 | Sequelae of cerebral infarction |
| G64..11 | CVA - cerebral artery occlusion |
| G64..13 | Stroke due to cerebral arterial occlusion |
| G671.00 | Generalised ischaemic cerebrovascular disease NOS |
| G6W..00 | Cereb infarct due unsp occlus/stenos precerebr arteries |
| G6X..00 | Cerebrl infarctn due/unspcf occlusn or sten/cerebrl artrs |
| Gyu6300 | [X]Cerebrl infarctn due/unspcf occlusn or sten/cerebrl artrs |
| Gyu6400 | [X]Other cerebral infarction |
| Gyu6500 | [X]Occlusion and stenosis of other precerebral arteries |
| Gyu6600 | [X]Occlusion and stenosis of other cerebral arteries |
| Gyu6G00 | [X]Cereb infarct due unsp occlus/stenos precerebr arteries |

##### Hypertension

| Read code | Read Term |
| --- | --- |
| 14A2.00 | H/O: hypertension |
| 2126100 | Hypertension resolved |
| 212K.00 | Hypertension resolved |
| 9OI9.00 | Hypertens.monitor deleted |
| 1JD..00 | Suspected hypertension |
| 246M.00 | White coat hypertension |
| 662..12 | Hypertension monitoring |
| 6629 | Hypertension:follow-up default |
| 662H.00 | Hypertension treatm.stopped |
| 662P.00 | Hypertension monitoring |
| 8CR4.00 | Hypertension clinical management plan |
| 9N03.00 | Seen in hypertension clinic |
| 9N1y200 | Seen in hypertension clinic |
| 9N4L.00 | DNA - Did not attend hypertension clinic |
| 9OI..00 | Hypertension monitoring admin. |
| 9OI..11 | Hypertension clinic admin. |
| 9OI1.00 | Attends hypertension monitor. |
| 9OI2.00 | Refuses hypertension monitor. |
| 9OI3.00 | Hyperten.monitor offer default |
| 9OI4.00 | Hypertens.monitor.1st letter |
| 9OI5.00 | Hypertens.monitor 2nd letter |
| 9OI6.00 | Hypertens.monitor 3rd letter |
| 9OI7.00 | Hypertens.monitor verbal inv. |
| 9OI8.00 | Hypertens.monitor phone invite |
| 9OIA.00 | Hypertension monitor.chck done |
| 9OIA.11 | Hypertension monitored |
| 9OIZ.00 | Hypertens.monitoring admin.NOS |
| 9h3..00 | Exception reporting: hypertension quality indicators |
| 9h31.00 | Excepted from hypertension qual indicators: Patient unsuit |
| 9h32.00 | Excepted from hypertension qual indicators: Informed dissent |
| 6624 | Borderline hyperten:yearly obs |
| 6627 | Good hypertension control |
| 6628 | Poor hypertension control |
| 662F.00 | Hypertension treatm. started |
| 662G.00 | Hypertensive treatm.changed |
| 662O.00 | On treatment for hypertension |
| 662b.00 | Moderate hypertension control |
| 662c.00 | Hypertension six month review |
| 662d.00 | Hypertension annual review |
| 662r.00 | Trial withdrawal of antihypertensive therapy |
| 7Q01.00 | High cost hypertension drugs |
| 8B26.00 | Antihypertensive therapy |
| 8BL0.00 | Patient on maximal tolerated antihypertensive therapy |
| 8I3N.00 | Hypertension treatment refused |
| F404200 | Blind hypertensive eye |
| F421300 | Hypertensive retinopathy |
| G2...00 | Hypertensive disease |
| G2...11 | BP - hypertensive disease |
| G20..00 | Essential hypertension |
| G200.00 | Malignant essential hypertension |
| G201.00 | Benign essential hypertension |
| G202.00 | Systolic hypertension |
| G203.00 | Diastolic hypertension |
| G20z.00 | Essential hypertension NOS |
| G20z.11 | Hypertension NOS |
| G21..00 | Hypertensive heart disease |
| G210.00 | Malignant hypertensive heart disease |
| G210000 | Malignant hypertensive heart disease without CCF |
| G210100 | Malignant hypertensive heart disease with CCF |
| G211.00 | Benign hypertensive heart disease |
| G211000 | Benign hypertensive heart disease without CCF |
| G211100 | Benign hypertensive heart disease with CCF |
| G21z.00 | Hypertensive heart disease NOS |
| G21z000 | Hypertensive heart disease NOS without CCF |
| G21z011 | Cardiomegaly - hypertensive |
| G21z100 | Hypertensive heart disease NOS with CCF |
| G21zz00 | Hypertensive heart disease NOS |
| G22..00 | Hypertensive renal disease |
| G220.00 | Malignant hypertensive renal disease |
| G221.00 | Benign hypertensive renal disease |
| G222.00 | Hypertensive renal disease with renal failure |
| G22z.00 | Hypertensive renal disease NOS |
| G22z.11 | Renal hypertension |
| G23..00 | Hypertensive heart and renal disease |
| G230.00 | Malignant hypertensive heart and renal disease |
| G231.00 | Benign hypertensive heart and renal disease |
| G232.00 | Hypertensive heart&renal dis wth (congestive) heart failure |
| G233.00 | Hypertensive heart and renal disease with renal failure |
| G234.00 | Hyperten heart&renal dis+both(congestv)heart and renal fail |
| G23z.00 | Hypertensive heart and renal disease NOS |
| G2y..00 | Other specified hypertensive disease |
| G2z..00 | Hypertensive disease NOS |
| G672.00 | Hypertensive encephalopathy |
| G672.11 | Hypertensive crisis |
| Gyu2.00 | [X]Hypertensive diseases |
| L122.00 | Other pre-existing hypertension in preg/childbirth/puerp |
| L122000 | Other pre-existing hypertension in preg/childb/puerp unspec |
| L122100 | Other pre-existing hypertension in preg/childb/puerp - deliv |
| L122300 | Other pre-exist hypertension in preg/childb/puerp-not deliv |
| L122z00 | Other pre-existing hypertension in preg/childb/puerp NOS |
| L127.00 | Pre-eclampsia or eclampsia with pre-existing hypertension |
| L127z00 | Pre-eclampsia or eclampsia + pre-existing hypertension NOS |
| L128.00 | Pre-exist hypertension compl preg childbirth and puerperium |
| L128000 | Pre-exist hyperten heart dis compl preg childbth+puerperium |
| L128200 | Pre-exist 2ndry hypertens comp preg childbth and puerperium |
| TJC7.00 | Adverse reaction to other antihypertensives |
| TJC7z00 | Adverse reaction to antihypertensives NOS |
| U60C500 | [X]Oth antihyperten drug caus advers eff in therap use, NEC |
| U60C511 | [X] Adverse reaction to other antihypertensives |
| U60C51A | [X] Adverse reaction to antihypertensives NOS |
| 6146200 | Hypertension induced by oral contraceptive pill |
| G24..00 | Secondary hypertension |
| G240.00 | Secondary malignant hypertension |
| G240000 | Secondary malignant renovascular hypertension |
| G240z00 | Secondary malignant hypertension NOS |
| G241.00 | Secondary benign hypertension |
| G241000 | Secondary benign renovascular hypertension |
| G241z00 | Secondary benign hypertension NOS |
| G244.00 | Hypertension secondary to endocrine disorders |
| G24z.00 | Secondary hypertension NOS |
| G24z000 | Secondary renovascular hypertension NOS |
| G24z100 | Hypertension secondary to drug |
| G24zz00 | Secondary hypertension NOS |
| Gyu2100 | [X]Hypertension secondary to other renal disorders |

##### Heart Failure

| Read code | Read term |
| --- | --- |
| 14A6.00 | H/O: heart failure |
| 14AM.00 | H/O: Heart failure in last year |
| 1736 | Paroxysmal nocturnal dyspnoea |
| 1J60.00 | Suspected heart failure |
| 23E1.00 | O/E - pulmonary oedema |
| 388D.00 | New York Heart Assoc classification heart failure symptoms |
| 662T.00 | Congestive heart failure monitoring |
| 662f.00 | New York Heart Association classification - class I |
| 662g.00 | New York Heart Association classification - class II |
| 662h.00 | New York Heart Association classification - class III |
| 662i.00 | New York Heart Association classification - class IV |
| 679X.00 | Heart failure education |
| 8CL3.00 | Heart failure care plan discussed with patient |
| 8HBE.00 | Heart failure follow-up |
| 8HHz.00 | Referral to heart failure exercise programme |
| 8Hg8.00 | Discharge from practice nurse heart failure clinic |
| 8Hk0.00 | Referred to heart failure education group |
| 9N0k.00 | Seen in heart failure clinic |
| 9N2p.00 | Seen by community heart failure nurse |
| 9N4s.00 | Did not attend practice nurse heart failure clinic |
| 9N4w.00 | Did not attend heart failure clinic |
| 9N6T.00 | Referred by heart failure nurse specialist |
| 9On..00 | Left ventricular dysfunction monitoring administration |
| 9On0.00 | Left ventricular dysfunction monitoring first letter |
| 9On1.00 | Left ventricular dysfunction monitoring second letter |
| 9On2.00 | Left ventricular dysfunction monitoring third letter |
| 9On3.00 | Left ventricular dysfunction monitoring verbal invite |
| 9On4.00 | Left ventricular dysfunction monitoring telephone invite |
| 9Or..00 | Heart failure monitoring administration |
| 9Or1.00 | Heart failure monitoring telephone invite |
| 9Or2.00 | Heart failure monitoring verbal invite |
| 9Or3.00 | Heart failure monitoring first letter |
| 9Or4.00 | Heart failure monitoring second letter |
| 9Or5.00 | Heart failure monitoring third letter |
| 9h1..00 | Exception reporting: LVD quality indicators |
| 9h11.00 | Excepted from LVD quality indicators: Patient unsuitable |
| 9h12.00 | Excepted from LVD quality indicators: Informed dissent |
| 9hH..00 | Exception reporting: heart failure quality indicators |
| 9hH0.00 | Excepted heart failure quality indicators: Patient unsuitabl |
| 9hH1.00 | Excepted heart failure quality indicators: Informed dissent |
| G581.12 | Pulmonary oedema - acute |
| G58z.11 | Weak heart |
| H54..00 | Pulmonary congestion and hypostasis |
| H541.00 | Pulmonary congestion |
| H541000 | Chronic pulmonary oedema |
| H541z00 | Pulmonary oedema NOS |
| H54z.00 | Pulmonary congestion and hypostasis NOS |
| H584.00 | Acute pulmonary oedema unspecified |
| H584z00 | Acute pulmonary oedema NOS |
| ZRad.00 | New York Heart Assoc classification heart failure symptoms |
| G580400 | Congestive heart failure due to valvular disease |
| G210.00 | Malignant hypertensive heart disease |
| G210000 | Malignant hypertensive heart disease without CCF |
| G210100 | Malignant hypertensive heart disease with CCF |
| G211100 | Benign hypertensive heart disease with CCF |
| G21z100 | Hypertensive heart disease NOS with CCF |
| G230.00 | Malignant hypertensive heart and renal disease |
| G232.00 | Hypertensive heart&renal dis wth (congestive) heart failure |
| G234.00 | Hyperten heart&renal dis+both(congestv)heart and renal fail |
| G1yz100 | Rheumatic left ventricular failure |
| 1O1..00 | Heart failure confirmed |
| 662W.00 | Heart failure annual review |
| 662p.00 | Heart failure 6 month review |
| 8B29.00 | Cardiac failure therapy |
| 8H2S.00 | Admit heart failure emergency |
| 9Or0.00 | Heart failure review completed |
| G400.00 | Acute cor pulmonale |
| G41z.11 | Chronic cor pulmonale |
| G554000 | Congestive cardiomyopathy |
| G554011 | Congestive obstructive cardiomyopathy |
| G58..00 | Heart failure |
| G58..11 | Cardiac failure |
| G580.00 | Congestive heart failure |
| G580.11 | Congestive cardiac failure |
| G580.12 | Right heart failure |
| G580.13 | Right ventricular failure |
| G580.14 | Biventricular failure |
| G580000 | Acute congestive heart failure |
| G580100 | Chronic congestive heart failure |
| G580200 | Decompensated cardiac failure |
| G580300 | Compensated cardiac failure |
| G581.00 | Left ventricular failure |
| G581.11 | Asthma - cardiac |
| G581.13 | Impaired left ventricular function |
| G581000 | Acute left ventricular failure |
| G582.00 | Acute heart failure |
| G58z.00 | Heart failure NOS |
| G58z.12 | Cardiac failure NOS |
| G5yy900 | Left ventricular systolic dysfunction |
| G5yyA00 | Left ventricular diastolic dysfunction |
| R2y1000 | [D]Cardiorespiratory failure |
| Q48y100 | Congenital cardiac failure |

##### CKD

| Read code | Read term |
| --- | --- |
| 1Z10.00 | Chronic kidney disease stage 1 |
| 1Z17.00 | Chronic kidney disease stage 1 with proteinuria |
| 1Z17.11 | CKD stage 1 with proteinuria |
| 1Z18.00 | Chronic kidney disease stage 1 without proteinuria |
| 1Z11.00 | Chronic kidney disease stage 2 |
| 1Z19.00 | Chronic kidney disease stage 2 with proteinuria |
| 1Z19.11 | CKD stage 2 with proteinuria |
| 1Z1A.00 | Chronic kidney disease stage 2 without proteinuria |
| 1Z1A.11 | CKD stage 2 without proteinuria |
| 1Z12.00 | Chronic kidney disease stage 3 |
| 1Z15.00 | Chronic kidney disease stage 3A |
| 1Z16.00 | Chronic kidney disease stage 3B |
| 1Z1B.00 | Chronic kidney disease stage 3 with proteinuria |
| 1Z1B.11 | CKD stage 3 with proteinuria |
| 1Z1C.00 | Chronic kidney disease stage 3 without proteinuria |
| 1Z1C.11 | CKD stage 3 without proteinuria |
| 1Z1D.00 | Chronic kidney disease stage 3A with proteinuria |
| 1Z1D.11 | CKD stage 3A with proteinuria |
| 1Z1E.00 | Chronic kidney disease stage 3A without proteinuria |
| 1Z1E.11 | CKD stage 3A without proteinuria |
| 1Z1F.00 | Chronic kidney disease stage 3B with proteinuria |
| 1Z1F.11 | CKD stage 3B with proteinuria |
| 1Z1G.00 | Chronic kidney disease stage 3B without proteinuria |
| 1Z13.00 | Chronic kidney disease stage 4 |
| 1Z1H.00 | Chronic kidney disease stage 4 with proteinuria |
| 1Z1J.00 | Chronic kidney disease stage 4 without proteinuria |
| 1Z1J.11 | CKD stage 4 without proteinuria |
| 1Z14.00 | Chronic kidney disease stage 5 |
| 1Z1K.00 | Chronic kidney disease stage 5 with proteinuria |
| 1Z1L.00 | Chronic kidney disease stage 5 without proteinuria |
| 1Z1L.11 | CKD stage 5 without proteinuria |

#### Non-Insulin Antidiabetic Drugs

##### Metformin

Glucophage 500mg tablets (Merck Serono Ltd)

Glucophage 850mg tablets (Merck Serono Ltd)

Glucophage Tablets 1000 mg

Glucophage SR 500mg tablets (Merck Serono Ltd)

Glucophage SR 750mg tablets (Merck Serono Ltd)

Glucophage SR 1000mg tablets (Merck Serono Ltd)

Metformin 500mg tablets

Metformin 850mg tablets

Bolamyn SR 1000mg tablets (Teva UK Ltd)

Metformin 500mg modified-release tablets

Metformin 1g modified-release tablets

Bolamyn SR 500mg tablets (Teva UK Ltd)

Metabet SR 500mg tablets (Morningside Healthcare Ltd)

Metabet SR 1000mg tablets (Morningside Healthcare Ltd)

Glucient SR 500mg tablets (Consilient Health Ltd)

Diagemet XL 500mg tablets (Genus Pharmaceuticals Ltd)

Glucient SR 750mg tablets (Consilient Health Ltd)

Glucient SR 1000mg tablets (Consilient Health Ltd)

Sukkarto SR 500mg tablets (Morningside Healthcare Ltd)

Sukkarto SR 1000mg tablets (Morningside Healthcare Ltd)

##### Sulfonylureas

Amaryl 2mg tablets (Zentiva)

Amaryl 1mg tablets (Zentiva)

Amaryl 3mg tablets (Zentiva)

Glimepiride 2mg tablets

Glimepiride 1mg tablets

Glimepiride 3mg tablets

Glimepiride 4mg tablets

Diamicron 80mg tablets (Servier Laboratories Ltd)

Gliclazide 80mg tablets

Gliclazide 30mg modified-release tablets

Diamicron 30mg MR tablets (Servier Laboratories Ltd)

Nazdol MR 30mg tablets (Consilient Health Ltd)

Dacadis MR 30mg tablets (Mylan Ltd)

Gliclazide 40mg tablets

Zicron 40mg tablets (Bristol Laboratories Ltd)

Edicil MR 30mg tablets (Teva UK Ltd)

Diamicron M/R tablets 60 mg

Gliclazide 60mg modified-release tablets

Laaglyda MR 60mg tablets (Consilient Health Ltd)

Vamju 30mg modified-release tablets (AMCo)

Vamju 60mg modified-release tablets (AMCo)

Bilxona 30mg modified-release tablets (Actavis UK Ltd)

Bilxona 60mg modified-release tablets (Actavis UK Ltd)

Glibenclamide 2.5mg tablets

Glibenclamide 5mg tablets

Tolbutamide 500mg tablets

Glipizide 5mg tablets

Minodiab 5mg tablets (Pfizer Ltd)

##### Other Oral Antidiabetic Drugs

Prandin 0.5mg tablets (Novo Nordisk Ltd)

Prandin 1mg tablets (Novo Nordisk Ltd)

Prandin 2mg tablets (Novo Nordisk Ltd)

Enyglid 0.5mg tablets (Consilient Health Ltd)

Enyglid 1mg tablets (Consilient Health Ltd)

Enyglid 2mg tablets (Consilient Health Ltd)

Repaglinide 1mg tablets

Repaglinide 2mg tablets

Repaglinide 500microgram tablets

Starlix 60mg tablets (Novartis Pharmaceuticals UK Ltd)

Starlix 120mg tablets (Novartis Pharmaceuticals UK Ltd)

Starlix 180mg tablets (Novartis Pharmaceuticals UK Ltd)

Trajenta 5mg tablets (Boehringer Ingelheim Ltd)

Alogliptin 6.25mg tablets

Alogliptin 12.5mg tablets

Alogliptin 25mg tablets

Vipidia 6.25mg tablets (Takeda UK Ltd)

Vipidia 12.5mg tablets (Takeda UK Ltd)

Vipidia 25mg tablets (Takeda UK Ltd)

Onglyza 5mg tablets (AstraZeneca UK Ltd)

Onglyza 2.5mg tablets (AstraZeneca UK Ltd)

Januvia 100mg tablets (Merck Sharp & Dohme Ltd)

Januvia 50mg tablets (Merck Sharp & Dohme Ltd)

Januvia 25mg tablets (Merck Sharp & Dohme Ltd)

Galvus 50mg tablets (Novartis Pharmaceuticals UK Ltd)

Canagliflozin 100mg tablets

Canagliflozin 300mg tablets

Invokana 100mg tablets (Janssen-Cilag Ltd)

Invokana 300mg tablets (Janssen-Cilag Ltd)

Forxiga 5mg tablets (AstraZeneca UK Ltd)

Forxiga 10mg tablets (AstraZeneca UK Ltd)

Jardiance 10mg tablets (Boehringer Ingelheim Ltd)

Jardiance 25mg tablets (Boehringer Ingelheim Ltd)

Actos 15mg tablets (Takeda UK Ltd)

Diabiom 15mg tablets (Tillomed Laboratories Ltd)

Actos 30mg tablets (Takeda UK Ltd)

Diabiom 30mg tablets (Tillomed Laboratories Ltd)

Glidipion 15mg tablets (Actavis UK Ltd)

Glidipion 30mg tablets (Actavis UK Ltd)

Glidipion 45mg tablets (Actavis UK Ltd)

Actos 45mg tablets (Takeda UK Ltd)

Diabiom 45mg tablets (Tillomed Laboratories Ltd)

Pioglitazone 15mg tablets

Pioglitazone 30mg tablets

Pioglitazone 45mg tablets

Acarbose 100mg tablets

Acarbose 50mg tablets

Glucobay 100mg tablets (Bayer Plc)

Glucobay 50mg tablets (Bayer Plc)

Alogliptin 12.5mg / Metformin 1g tablets

Vipdomet 12.5mg/1000mg tablets (Takeda UK Ltd)

Jentadueto 2.5mg/1000mg tablets (Boehringer Ingelheim Ltd)

Jentadueto 2.5mg/850mg tablets (Boehringer Ingelheim Ltd)

Komboglyze 2.5mg/850mg tablets (AstraZeneca UK Ltd)

Komboglyze 2.5mg/1000mg tablets (AstraZeneca UK Ltd)

Janumet 50mg/1000mg tablets (Merck Sharp & Dohme Ltd)

Eucreas 50mg/1000mg tablets (Novartis Pharmaceuticals UK Ltd)

Eucreas 50mg/850mg tablets (Novartis Pharmaceuticals UK Ltd)

Vokanamet 50mg/850mg tablets (Janssen-Cilag Ltd)

Vokanamet 50mg/1000mg tablets (Janssen-Cilag Ltd)

Xigduo 5mg/1000mg tablets (AstraZeneca UK Ltd)

Xigduo 5mg/850mg tablets (AstraZeneca UK Ltd)

Competact 15mg/850mg tablets (Takeda UK Ltd)

Byetta 5micrograms/0.02ml solution for injection 1.2ml pre-filled disposable devices (AstraZeneca UK Ltd)

Byetta 10micrograms/0.04ml solution for injection 2.4ml pre-filled disposable devices (AstraZeneca UK Ltd)

Bydureon 2mg powder and solvent for suspension for prolonged-release injection vials (AstraZeneca UK Ltd)

Bydureon 2mg powder and solvent for prolonged-release suspension for injection pre-filled pen (AstraZeneca UK Ltd)

Liraglutide 6mg/ml solution for injection 3ml pre-filled disposable devices

Victoza 6mg/ml solution for injection 3ml pre-filled pen (Novo Nordisk Ltd)

Lyxumia 10micrograms/0.2ml solution for injection 3ml pre-filled pen (Sanofi)

Lyxumia 20micrograms/0.2ml solution for injection 3ml pre-filled pen (Sanofi)

#### Insulin

NovoRapid 100units/ml solution for injection 10ml vials (Novo Nordisk Ltd)

NovoRapid FlexPen 100units/ml solution for injection 3ml pre-filled pen (Novo Nordisk Ltd)

NovoRapid FlexTouch 100units/ml solution for injection 3ml pre-filled pen (Novo Nordisk Ltd)

NovoRapid Penfill 100units/ml solution for injection 3ml cartridges (Novo Nordisk Ltd)

NovoRapid PumpCart 100units/ml solution for injection 1.6ml cartridges (Novo Nordisk Ltd)

Apidra 100units/ml solution for injection 10ml vials (Sanofi)

Apidra 100units/ml solution for injection 3ml cartridges (Sanofi)

Apidra 100units/ml solution for injection 3ml pre-filled SoloStar pen (Sanofi)

Humalog 100units/ml solution for injection 10ml vials (Eli Lilly and Company Ltd)

Humalog 100units/ml solution for injection 3ml cartridges (Eli Lilly and Company Ltd)

Humalog KwikPen 100units/ml solution for injection 3ml pre-filled pen (Eli Lilly and Company Ltd)

Humalog KwikPen 200units/ml solution for injection 3ml pre-filled pen (Eli Lilly and Company Ltd)

Actrapid 100units/ml solution for injection 10ml vials (Novo Nordisk Ltd)

Humulin R Injection 100 units/ml, 10 ml vial

Humulin S 100units/ml solution for injection 10ml vials (Eli Lilly and Company Ltd)

Humulin S 100units/ml solution for injection 3ml cartridges (Eli Lilly and Company Ltd)

Hypurin Bovine Neutral 100units/ml solution for injection 10ml vials (Wockhardt UK Ltd)

Hypurin Bovine Neutral 100units/ml solution for injection 3ml cartridges (Wockhardt UK Ltd)

Hypurin Porcine Neutral 100units/ml solution for injection 3ml cartridges (Wockhardt UK Ltd)

Hypurin Porcine Neutral 100units/ml solution for injection 10ml vials (Wockhardt UK Ltd)

Insuman Infusat 100units/ml solution for injection 10ml vials (Sanofi)

Insuman Infusat 100units/ml solution for injection 3.15ml cartridges (Sanofi)

Insuman Rapid 100units/ml solution for injection 3ml cartridges (Sanofi)

Humulin R 500units/ml solution for injection 20ml vials (Imported (United States))

Humulin I 100units/ml suspension for injection 10ml vials (Eli Lilly and Company Ltd)

Humulin I 100units/ml suspension for injection 3ml cartridges (Eli Lilly and Company Ltd)

Humulin I KwikPen 100units/ml suspension for injection 3ml pre-filled pen (Eli Lilly and Company Ltd)

Hypurin Bovine Isophane 100units/ml suspension for injection 10ml vials (Wockhardt UK Ltd)

Hypurin Bovine Isophane 100units/ml suspension for injection 3ml cartridges (Wockhardt UK Ltd)

Hypurin Porcine Isophane 100units/ml suspension for injection 3ml cartridges (Wockhardt UK Ltd)

Hypurin Porcine Isophane 100units/ml suspension for injection 10ml vials (Wockhardt UK Ltd)

Insulatard 100units/ml suspension for injection 10ml vials (Novo Nordisk Ltd)

Insulatard InnoLet 100units/ml suspension for injection 3ml pre-filled pen (Novo Nordisk Ltd)

Insulatard Penfill 100units/ml suspension for injection 3ml cartridges (Novo Nordisk Ltd)

Insuman Basal 100units/ml suspension for injection 5ml vials (Sanofi)

Insuman Basal 100units/ml suspension for injection 3ml cartridges (Sanofi)

Insuman Basal 100units/ml suspension for injection 3ml pre-filled SoloStar pen (Sanofi)

Humulin M3 100units/ml suspension for injection 10ml vials (Eli Lilly and Company Ltd)

Humulin M3 100units/ml suspension for injection 3ml cartridges (Eli Lilly and Company Ltd)

Humulin M3 KwikPen 100units/ml suspension for injection 3ml pre-filled pen (Eli Lilly and Company Ltd)

Hypurin Porcine 30/70 Mix 100units/ml suspension for injection 10ml vials (Wockhardt UK Ltd)

Hypurin Porcine 30/70 Mix 100units/ml suspension for injection 3ml cartridges (Wockhardt UK Ltd)

Insuman Comb 15 100units/ml suspension for injection 3ml cartridges (Sanofi)

Insuman Comb 25 100units/ml suspension for injection 3ml cartridges (Sanofi)

Insuman Comb 25 100units/ml suspension for injection 3ml pre-filled SoloStar pen (Sanofi)

Insuman Comb 25 100units/ml suspension for injection 5ml vials (Sanofi)

Insuman Comb 50 100units/ml suspension for injection 3ml cartridges (Sanofi)

NovoMix 30 FlexPen 100units/ml suspension for injection 3ml pre-filled pen (Novo Nordisk Ltd)

NovoMix 30 Penfill 100units/ml suspension for injection 3ml cartridges (Novo Nordisk Ltd)

Humalog Mix25 100units/ml suspension for injection 3ml cartridges (Eli Lilly and Company Ltd)

Humalog Mix25 100units/ml suspension for injection 10ml vials (Eli Lilly and Company Ltd)

Humalog Mix25 KwikPen 100units/ml suspension for injection 3ml pre-filled pen (Eli Lilly and Company Ltd)

Humalog Mix50 100units/ml suspension for injection 3ml cartridges (Eli Lilly and Company Ltd)

Humalog Mix50 KwikPen 100units/ml suspension for injection 3ml pre-filled pen (Eli Lilly and Company Ltd)

Hypurin Bovine Lente 100units/ml suspension for injection 10ml vials (Wockhardt UK Ltd)

Hypurin Bovine Protamine Zinc 100units/ml suspension for injection 10ml vials (Wockhardt UK Ltd)

Abasaglar 100units/ml solution for injection 3ml cartridges (Eli Lilly and Company Ltd)

Abasaglar KwikPen 100units/ml solution for injection 3ml pre-filled pen (Eli Lilly and Company Ltd)

Lantus 100units/ml solution for injection 10ml vials (Sanofi)

Lantus 100units/ml solution for injection 3ml cartridges (Sanofi)

Lantus 100units/ml solution for injection 3ml pre-filled SoloStar pen (Sanofi)

Toujeo 300units/ml solution for injection 1.5ml pre-filled SoloStar pen (Sanofi)

Levemir FlexPen 100units/ml solution for injection 3ml pre-filled pen (Novo Nordisk Ltd)

Levemir InnoLet 100units/ml solution for injection 3ml pre-filled pen (Novo Nordisk Ltd)

Levemir Penfill 100units/ml solution for injection 3ml cartridges (Novo Nordisk Ltd)

Tresiba FlexTouch 100units/ml solution for injection 3ml pre-filled pen (Novo Nordisk Ltd)

Tresiba FlexTouch 200units/ml solution for injection 3ml pre-filled pen (Novo Nordisk Ltd)

Tresiba Penfill 100units/ml solution for injection 3ml cartridges (Novo Nordisk Ltd)

Xultophy 100units/ml / 3.6mg/ml solution for injection 3ml pre-filled pen (Novo Nordisk Ltd)
